# Prevalence and risk/protective indicators of peri-implant diseases: a university-representative cross-sectional study

**DOI:** 10.1101/2020.06.04.20121970

**Authors:** Mario Romandini, Cristina Lima, Ignacio Pedrinaci, Ana Araoz, Maria Costanza Soldini, Mariano Sanz

## Abstract

**Aim:** To evaluate the prevalence of peri-implant diseases and to identify risk/protective indicators of peri-implantitis.

**Materials and Methods:** 240 randomly selected patients from a university clinic database were invited to participate. Those who accepted, once data from their medical and dental history was collected, were examined clinically and radiographically to assess the prevalence of peri-implant health and diseases. A multilevel multivariate logistic regression analysis was carried out to identify those factors associated either positively (risk) or negatively (protective) with peri-implantitis defined as BoP/SoP and bone levels ≧2 mm.

**Results:** 99 patients with a total of 458 dental implants were analyzed. The prevalence of preperiimplantitis and of peri-implantitis were respectively 56.6% and 31.3% at patient-level, while 27.9% and 31.7% at implant-level. The following factors were identified as risk indicators for periimplantitis: smoking (OR = 3.59; 95%CI:1.52–8.45), moderate/severe periodontitis (OR = 2.77; 95%CI:1.20–6.36), < 16 remaining teeth (OR = 2.23; 95%CI:1.05–4.73), plaque (OR = 3.49; 95%CI:1.13–10.75), implant malposition (too vestibular: OR = 2.85; 95%CI:1.17–6.93), implant brand (Nobel vs. Straumann: OR = 4.41;95% CI:1.76–11.09), restoration type (bridge: OR = 2.47; 95%CI:1.19–5.12), and trauma as reason of tooth loss (OR = 6.51;95% CI:1.45–29.26). Conversely, the following factors were identified as protective indicators: interproximal flossing/brushing (OR = 0.27; 95%CI:0.11–0.68), proton pump inhibitors (OR = 0.08; 95%CI:0.01–0.90) and anticoagulants (OR = 0.08; 95%CI:0.01–0.56).

**Conclusions:** Peri-implant diseases are highly prevalent among patients with dental implants in this university-based population. Several factors were identified as risk- and protective-indicators of peri-implantitis.

## Introduction

Peri-implantitis has been defined in the 2017 World Workshop as a plaque-associated pathological condition affecting tissues around dental implants, characterized by inflammation in the peri-implant mucosa and subsequent progressive loss of supporting bone (Berglundh et al., 2018a; Schwarz, Derks, Monje, & Wang, 2018). Peri-implantitis has shown a high prevalence among patients with dental implants for extended periods of time (Derks & Tomasi, 2015; Derks, Schaller, Hakansson, Wennstrom, Tomasi, & Berglundh, 2016a; Rakic et al., 2018; Rodrigo et al., 2018; Romandini, Cordaro, Donno, & Cordaro, 2019; Vignoletti, Di Domenico, Di Martino, Montero, & De Sanctis, 2019; Wada et al., 2019) and, when proper therapy is not provided, its progression follows a non-linear and accelerating pattern, which can ultimately result in implant loss (Derks, Schaller, Håkansson, et al., 2016b).

Despite different non-surgical and surgical treatment strategies proposed for treating periimplantitis, disease resolution is seldom the long-term outcome and even when achieved, recurrence may occur (Berglundh, Wennström, & Lindhe, 2018b; Carcuac et al., 2017; Cha, Lee, & Kim, 2019; de Tapia et al., 2019; Figuero, Graziani, Sanz, Herrera, & Sanz, 2014; Heitz-Mayfield et al., 2018; Nart et al., 2020; Ravidà, Saleh, et al., 2020b; Roccuzzo, Layton, Roccuzzo, & Heitz-Mayfield, 2018). In light of this limited predictability, its prevention becomes of uttermost importance. The main strategy for preventing peri-implantitis is the management of peri-implant mucositis (Barootchi, Ravidà, Tavelli, & Wang, 2020; Jepsen et al., 2015), since this condition is its reversible precursor (Berglundh et al., 2018a; Costa et al., 2012). This approach should be combined with the control of modifiable risk factors, although presently there is a limited number of true risk factors of peri-implantitis demonstrated in prospective cohort investigations (Heitz-Mayfield & Salvi, 2018; Heitz-Mayfield, Heitz, & Lang, 2020; Schwarz, Derks, Monje, & Wang, 2018). Consequently, a fundamental thrive in current implant research is the identification of further risk factors and indicators of peri-implant diseases.

Although many epidemiological studies have reported data on the prevalence and risk indicators of peri-implant diseases, there are only two epidemiological studies that have used representative samples (Derks, Schaller, Hakansson, Wennstrom, Tomasi, & Berglundh, 2016a; Rodrigo et al., 2018), both of them referring to private settings and not to university clinics. Therefore, there is still a need of further studies in different settings and with representative samples, since the use of convenience samples (e.g. maintenance patients) may hamper the true analysis of the disease prevalence and risk indicators (Sanz & Chapple, 2012). It was, therefore, the aim of this cross-sectional study to analyze in a university-representative sample, the prevalence of peri-implant diseases as well as to study the risk/protective indicators of peri-implantitis.

## Materials and Methods

The present cross-sectional study is reported according to the STrengthening the Reporting of OBservational studies in Epidemiology (STROBE) guidelines (Elm et al., 2007; Vandenbroucke et al., 2007). The research protocol was ethically approved by the CEIC Hospital Clínico San Carlos, Madrid, Spain (19/182-E).

## Sampling procedures

The sample size calculation was based on the null hypothesis that the prevalence of peri-implantitis in the present sample was the same as 45.0% in Derks et al. (2016a). For a 10% threshold in prevalence difference and with an alpha level set at 0.05, a sample of 96 participants would result in 80% power to reject the null hypothesis. Considering that this study was carried out on a representative sample, it was reasonable to select at least 200 participants, as it was expected a high declination rate (about 50%) of those invited to participate (e.g. due to change of city or telephone number, or death).

To generalize the results to all the patients who received implants in the Master of Periodontology of the Complutense University of Madrid, we used a complex protocol through a stratified multistage sampling (Sanz & Chapple, 2012; Tomasi & Derks, 2012). Three patients were randomly selected, by computer-generated randomization lists, from each periodontist (or postgraduate student in periodontology) that placed implants in at least 10 patients, from September 2000 to July 2017, in the referred clinic. During each academic year, periodontists who placed implants in less than 10 patients were grouped in a single category and 3 patients were also selected for this group.

The selected patients were invited to participate in the study by telephone calls on the numbers reported in their clinical charts, and if no response, the patient was not discarded until at least five attempts on different days have been made. Only implants with at least one year of loading were evaluated.

## Data collection

The participants who accepted to participate underwent a through data collection process, consisting of four phases: collection of demographic and medical/dental history data, a clinical examination, a radiographic examination and an analysis of their past dental records. Its detailed description is reported in the Appendix.

Briefly, the history collection was structured in two steps. The first one (self-reported questionnaire) was based on the completion of written questionnaires by the study participants, after a brief explanation by one interviewer. The second step (structured interview) was based on a series of standardized questions asked by a trained interviewer.

The clinical oral examination was carried out by 2 calibrated examiners and included the assessment of patient-, restoration- and implant- related variables. Regarding patient-related variables, they included the assessment of periodontal status according to the AAP/CDC case definitions (Eke, Page, Wei, Thornton-Evans, & Genco, 2012) and the number of remaining teeth.

The implant supported restoration data included the type of restoration and its retention. The implant-related examination included the assessment of the location of each implant, its correct placement (adequate or mispositioned), the presence of adjacent teeth, keratinized tissue height (KTH), mobility of mucosal margin, biotype, tissue thickness, clinical signs of occlusal overloading on implants and of bruxism (Carra, Huynh, & Lavigne, 2012), and presence of a prosthetic design not allowing access to hygiene. Each implant was examined using a manual UNC-15 periodontal probe (PCP15; Hu-Friedy, Chicago, IL, USA) at 6 sites/implant for the following measurements: presence of visible plaque, recession depth (Sanz-Martín et al., 2020), probing pocket depth (PPD), bleeding and suppuration on probing (BoP/SoP, within 30s). The two examiners were calibrated at the start of the study to apply the same examination criteria for each item and the inter-rater agreement on 10 patients was also calculated (Appendix).

Peri-apical digital radiographs of the included implants were obtained. One calibrated investigator measured the marginal bone level (BL) from the implant shoulder to the first bone-implant contact, using a software program (Autocad 2016 TM, AutoDesk Inc., San Rafael, CA, USA) (Flores-Guillen, Álvarez-Novoa, Barbieri, Martin, & Sanz, 2018). One month after the initial evaluation, 50 randomly selected radiographs were re-measured by the same investigator to calculate the intraexaminer agreement (ICC = 0.98; 95% CI 0.96–0.99; p< 0.001).

Original dental charts for each of the included implants were also analyzed to extract data on the implant brand and the implant dimensions (length, width and eventual collar length). If the original information was not available (i.e. lost dental chart or implant placed outside the clinic), we attempted to deduce the corresponding information from the radiographs (Appendix). A validation of this method was performed on thirty randomly selected implants of known dimensions, which resulted in an ICC for implant length of 0.95 (95% CI 0.89–0.97; p< 0.001).

## Peri-implant health and diseases case definitions

The following case definitions were used (Sanz & Chapple, 2012):

- Peri-implant health: absence of BoP/SoP;
- Peri-implant mucositis: presence of BoP/SoP together with radiographic BL< 1 mm;
- Pre-periimplantitis: presence of BoP/SoP together with 1 mm≦BL<2 mm
- Peri-implantitis: presence of BoP/SoP together with radiographic BL ≧ 2 mm.

Also, in order to facilitate the comparison of the present findings with other studies, peri-implantitis was further reported with a BL≧3 mm as severity cut-off of bone levels (severe peri-implantitis), which corresponds to the 2017 World Workshop classification (Berglundh et al., 2018a) case definition for epidemiological studies.

## Data analysis

All statistical analyses were performed with STATA version 13.1 software (StataCorp LP, College Station, TX, USA). Descriptive characteristics regarding all the covariates were summarized. Periimplant health and diseases prevalence were calculated both at implant- and at patient-level. In addition to reporting its prevalence with the ≥3 mm threshold, peri-implantitis severity was expressed as the mean bone levels in implants with peri-implantitis and as the percentage of bone levels in relation to the implant length in implants with peri-implantitis.

The extent of peri-implantitis was assessed in patients with > 1 implant, as the mean number and the percentage of affected implants in subjects with peri-implantitis.

Risk/protective indicators for peri-implantitis were studied using multilevel multivariate logistic regression analyses (patient- and implant-level). Due to the paucity of information on true risk factors available in literature, an exploratory approach was used. Each potential indicator was tested individually by adding it to an empty model having as dependent variable the peri-implantitis status and testing the significance. All variables that were significant at the 0.10 level were included in an intermediate multivariate model and non-significant variables were sequentially removed. The final model included all factors that remained significant (p< 0.05).

## Results

The sampling strategy resulted in the selection of 240 subjects and 110 of them accepted to participate receiving the examination. From this initial sample, one patient was excluded as only presenting one implant loaded from less than 1 year, while another patient was excluded due to the loss of all the implants. Due to the absence of readable radiographs of all the implants, 9 further patients were excluded from the present analysis, resulting in a total analyzed sample of 99 patients. Those 99 patients had a total of 475 implants; however, 2 implants were excluded since were loaded from less than 1 year and 15 of them due to the absence of readable radiographs. Consequently, the present analysis included a total of 99 patients with 458 dental implants with at least 1 year of loading time.

## Descriptive statistics of the study population and implants

Table 1 and table 2 provides descriptive statistics of the study population and implants. Most of the included patients were women (60.61%), currently non-smokers (81.81%), with moderate/severe periodontitis (62.89%) and with a mean age at examination of 63.71 years. Most of the implants were placed in the maxilla (55.24%) and rehabilitated through bridges (58.30%) and screw-retained prosthesis (49.34%). The distribution of the potential risk/protective indicators in the study population and implants according to their peri-implant status is reported in tables S1 and S2.

**Table 1.** General characteristics of the study population.

|  | **N=99** |
| --- | --- |
| **Age** (years), mean (SD) | 63.71 (9.26) |
| **Gender**, N (%) |  |
| Male | 39 (39.39) |
| Female | 60 (60.61) |
| **Educational Level**, N (%) |  |
| Primary school | 32 (32.32) |
| High school | 26 (26.26) |
| Middle grade | 20 (20.21) |
| University/College | 21 (21.21) |
| **Smoking Status**, N (%) |  |
| Non-smokers | 41 (41.41) |
| Former smokers | 40 (40.40) |
| Current smokers | 18 (18.18) |
| **Marital Status**, N (%) |  |
| Married | 73 (73.74) |
| Widow | 6 (6.06) |
| Divorced | 9 (9.09) |
| Never married | 8 (8.08) |
| Living with unmarried partner | 3 (3.03) |
| **BMI** (kg/m<sup>2</sup>), mean (SD) | 25.55 (3.66) |
| **Diabetes Status**, N (%) |  |
| No diabetes | 83 (83.84) |
| Diabetes | 16 (16.16) |
| **Periodontal Status (AAP)**, N (%) |  |
| No/Mild Periodontitis | 27 (27.84) |
| Moderate/Severe Periodontitis | 61 (62.89) |
| Edentulous | 9 (9.28) |
| **Regular Maintenance**, N (%) |  |
| No | 46 (46.94) |
| Less than one/year | 8 (8.16) |
| One/year | 21 (21.43) |
| Two/year | 19 (19.39) |
| Three or more/year | 4 (4.08) |

Total number varies according to missing data for each variable.
SD, standard deviation; N, number.

**Table 2.** General characteristics of the study implants.

|  | N=458 |
| --- | --- |
| **Jaw, N (%)** |  |
| Maxilla | 253 (55.2) |
| Mandible | 205 (44.8) |
| **Position, N (%)** |  |
| Anterior (canine-canine) | 83 (18.1) |
| Posterior | 375 (81.9) |
| **Side, N (%)** |  |
| Right | 234 (51.1) |
| Left | 224 (48.9) |
| **Type of Prosthesis, N (%)** |  |
| Single crown | 136 (29.7) |
| Bridge | 267 (58.3) |
| Overdenture | 14 (3.1) |
| Full-arch fixed restoration | 41 (8.9) |
| **Prosthesis Retention, N (%)** |  |
| Cemented | 218 (47.6) |
| Screw-retained | 226 (49.3) |
| Locator | 8 (1.8) |
| Bar | 6 (1.3) |
| **Reason of Tooth Loss, N (%)** |  |
| Caries | 185 (40.4) |
| Periodontitis | 151 (32.9) |
| Trauma | 15 (3.3) |
| Agnesia | 8 (1.8) |
| Other reason/Unknown | 99 (21.6) |
| **Implant Brand, N (%)** |  |
| S | 230 (50.7) |
| N | 57 (12.6) |
| A | 76 (16.7) |
| Other | 91 (20.0) |
| **Implant Length** (mm), mean (SD) | 9.9 (1.7) |
| **Implant Diameter** (mm), mean (SD) | 4.1 (0.4) |

Total number varies according to missing data for each variable.
Implant brands: S, Straumann; N, Nobel Biocare; A, AstraTech.
SD, standard deviation; N, number.

## Prevalence and severity of peri-implant health and diseases

The prevalence of peri-implant health and diseases is reported in table 3.

**Table 3.** Prevalence of peri-implant health and diseases.

|  | **Patient-level** |  | **Implant-level** |  |
| --- | --- | --- | --- | --- |
|  | *Inflammation cut-off: BoP/SoP at least 1/6 site* | *Inflammation cut-off: BoP/SoP at least 4/6 sites* | *Inflammation cut-off: BoP/SoP at least 1/6 site* | *Inflammation cut-off: BoP/SoP at least 4/6 sites* |
| **Peri-implant health, N (%)** | 1 (1.0) | 27 (27.3) | 39 (8.5) | 253 (55.2) |
| **Peri-implant mucositis, N (%)** | 11 (11.1) | 18 (18.2) | 146 (31.9) | 82 (17.9) |
| **Pre-Periimplantitis, N (%)** | 31 (31.3) | 23 (23.2) | 109 (31.7) | 65 (14.2) |
| **Peri-implantitis, N (%)** |  |  |  |  |
| *BL* $\geq 2$ mm † | 56 (56.6) | 31 (31.3) | 128 (27.9) | 58 (12.7) |
| *Severe (BL* $\geq 3$ mm) ‡ | 23 (23.2) | 12 (12.1) | 57 (12.4) | 26 (5.7) |

N, number; BoP, bleeding on probing; SoP, suppuration on probing; BL, bone levels at the examination.
† Corresponding to the Sanz & Chapple (2012) peri-implantitis case definition.
‡ Corresponding to the 2017 World Workshop peri-implantitis case definition for epidemiological studies.

**Table 4.**
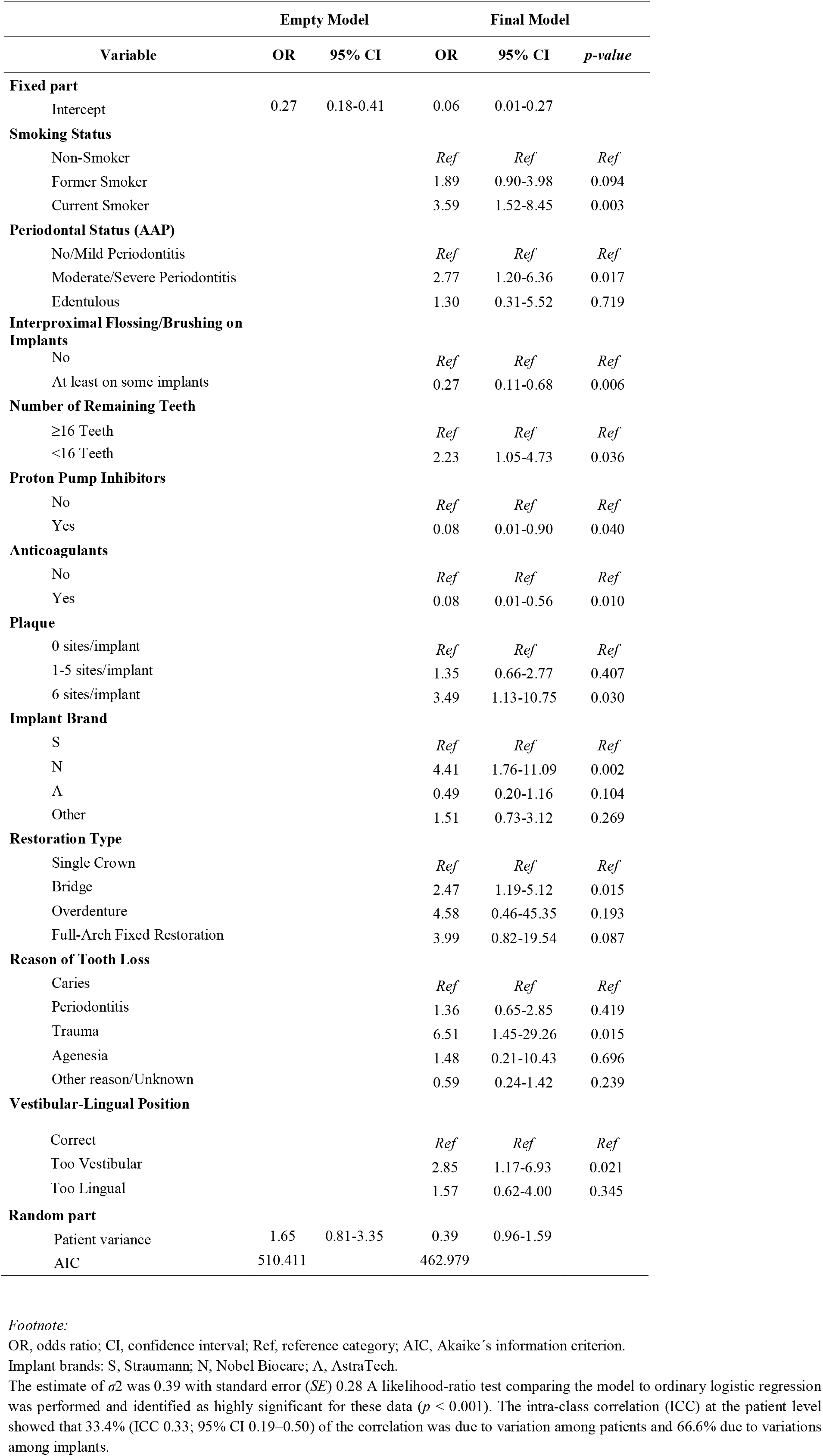
Risk/protective indicators associated with peri-implantitis: multilevel multivariate logistic regression analysis.

| Variable | Empty Model |  | Final Model |  |  |
| --- | --- | --- | --- | --- | --- |
|  | OR | 95% CI | OR | 95% CI | *p-value* |
| **Fixed part** |  |  |  |  |  |
| Intercept | 0.27 | 0.18-0.41 | 0.06 | 0.01-0.27 |  |
| **Smoking Status** |  |  |  |  |  |
| Non-Smoker |  |  | *Ref* | *Ref* | *Ref* |
| Former Smoker |  |  | 1.89 | 0.90-3.98 | 0.094 |
| Current Smoker |  |  | 3.59 | 1.52-8.45 | 0.003 |
| **Periodontal Status (AAP)** |  |  |  |  |  |
| No/Mild Periodontitis |  |  | *Ref* | *Ref* | *Ref* |
| Moderate/Severe Periodontitis |  |  | 2.77 | 1.20-6.36 | 0.017 |
| Edentulous |  |  | 1.30 | 0.31-5.52 | 0.719 |
| **Interproximal Flossing/Brushing on Implants** |  |  |  |  |  |
| No |  |  | *Ref* | *Ref* | *Ref* |
| At least on some implants |  |  | 0.27 | 0.11-0.68 | 0.006 |
| **Number of Remaining Teeth** |  |  |  |  |  |
| ≥16 Teeth |  |  | *Ref* | *Ref* | *Ref* |
| <16 Teeth |  |  | 2.23 | 1.05-4.73 | 0.036 |
| **Proton Pump Inhibitors** |  |  |  |  |  |
| No |  |  | *Ref* | *Ref* | *Ref* |
| Yes |  |  | 0.08 | 0.01-0.90 | 0.040 |
| **Anticoagulants** |  |  |  |  |  |
| No |  |  | *Ref* | *Ref* | *Ref* |
| Yes |  |  | 0.08 | 0.01-0.56 | 0.010 |
| **Plaque** |  |  |  |  |  |
| 0 sites/implant |  |  | *Ref* | *Ref* | *Ref* |
| 1-5 sites/implant |  |  | 1.35 | 0.66-2.77 | 0.407 |
| 6 sites/implant |  |  | 3.49 | 1.13-10.75 | 0.030 |
| **Implant Brand** |  |  |  |  |  |
| S |  |  | *Ref* | *Ref* | *Ref* |
| N |  |  | 4.41 | 1.76-11.09 | 0.002 |
| A |  |  | 0.49 | 0.20-1.16 | 0.104 |
| Other |  |  | 1.51 | 0.73-3.12 | 0.269 |
| **Restoration Type** |  |  |  |  |  |
| Single Crown |  |  | *Ref* | *Ref* | *Ref* |
| Bridge |  |  | 2.47 | 1.19-5.12 | 0.015 |
| Overdenture |  |  | 4.58 | 0.46-45.35 | 0.193 |
| Full-Arch Fixed Restoration |  |  | 3.99 | 0.82-19.54 | 0.087 |
| **Reason of Tooth Loss** |  |  |  |  |  |
| Caries |  |  | *Ref* | *Ref* | *Ref* |
| Periodontitis |  |  | 1.36 | 0.65-2.85 | 0.419 |
| Trauma |  |  | 6.51 | 1.45-29.26 | 0.015 |
| Agnesia |  |  | 1.48 | 0.21-10.43 | 0.696 |
| Other reason/Unknown |  |  | 0.59 | 0.24-1.42 | 0.239 |
| **Vestibular-Lingual Position** |  |  |  |  |  |
| Correct |  |  | *Ref* | *Ref* | *Ref* |
| Too Vestibular |  |  | 2.85 | 1.17-6.93 | 0.021 |
| Too Lingual |  |  | 1.57 | 0.62-4.00 | 0.345 |
| **Random part** |  |  |  |  |  |
| Patient variance | 1.65 | 0.81-3.35 | 0.39 | 0.96-1.59 |  |
| AIC | 510.411 |  | 462.979 |  |  |

OR, odds ratio; CI, confidence interval; Ref, reference category; AIC, Akaike's information criterion.
Implant brands: S, Straumann; N, Nobel Biocare; A, AstraTech.
The estimate of $\sigma^2$ was 0.39 with standard error (*SE*) 0.28. A likelihood-ratio test comparing the model to ordinary logistic regression was performed and identified as highly significant for these data ( $p < 0.001$ ). The intra-class correlation (ICC) at the patient level showed that 33.4% (ICC 0.33; 95% CI 0.19–0.50) of the correlation was due to variation among patients and 66.6% due to variations among implants.

At patient-level, the prevalence of peri-implant health was 1.0%, of peri-implant mucositis of 11.1%, of pre-periimplantitis of 31.3% and of peri-implantitis of 56.6%. Severe peri-implantitis, corresponding to the case definition of the 2017 World Workshop classification, was present in 23.2% of the patients.

At implant-level, the prevalence of peri-implant health was 8.5%, of peri-implant mucositis of 31.9%, of pre-periimplantitis of 31.7% and of peri-implantitis of 27.9%. Severe peri-implantitis with BL≧3 mm was present in 12.4% of the study implants.

The mean bone levels in the 128 implants presenting with peri-implantitis was 3.15 (SD = 1.30), while for the 57 implants with severe peri-implantitis was 4.10 (SD = 1.45). The bone levels at implants with peri-implantitis corresponded to 32.21% (SD = 13.14) of the intraosseous portion of the implant, while for severe peri-implantitis to 41.97% (SD = 13.74).

When considering a threshold of at least 4/6 sites with BoP/SoP, the prevalences of peri-implantitis decreased, with a corresponding increase of peri-implant health cases (27.27% at patient-level and 55.24% at implant-level).

## Extent of peri-implantitis

Peri-implantitis was detected in 55 of 89 patients with > 1 implant. The mean number of implants in this category of patients was 5.20 ± 2.91 and the mean number of implants with peri-implantitis was 2.31 ± 1.71, which represented 43.57% (SD = 25.44) of their implants.

## Risk/protective indicators for peri-implantitis

In univariate analyses, peri-implantitis was associated (p< 0.1), either in a direct or inverse manner, with the following patient-level characteristics (table S3): smoking status, history of cardiovascular diseases, history of osteopenia/osteoporosis, periodontal status, history of periodontal treatment, interproximal flossing and/or brushing on implants, tooth-brushing frequency, number of remaining teeth, use of proton pump inhibitors (PPIs), use of anticoagulants, and use of insulin. Moreover, it was associated (p< 0.1) with the following implant-level variables (table S4): presence of plaque, keratinized tissue height, tissue thickness, peri-implant phenotype, vestibular-lingual position of the implant, implant brand, restoration type, restoration retention, reason of tooth loss, position of the replaced tooth, presence of an adjacent tooth, clinical signs of occlusal overloading, and presence of platform switching.

However, in the final multi-level multi-variate logistic regression model, only the following factors remained significant at the 0.05 level: smoking status (smokers vs. non-smokers: OR = 3.59; 95%CI:1.52–8.45), periodontal status (moderate/severe periodontitis vs. no/mild periodontitis: OR = 2.77; 95%CI:1.20–6.36), interproximal flossing/brushing on implants (OR = 0.27; 95%CI:0.11–0.68), number of remaining teeth (< 16 vs. ≧16: OR = 2.23; 95%CI:1.05–4.73), use of PPIs(OR = 0.08; 95%CI:0.01–0.90), use of anticoagulants (OR = 0.08; 95%CI:0.01–0.56), presence of plaque (6 sites/implant vs. 0 sites/implant: OR = 3.49; 95%CI:1.13–10.75), implant brand (N vs. S: OR = 4.41; 95%CI:1.76–11.09), restoration type (bridge vs. single crown: OR = 2.47; 95%CI:1.19–5.12), reason of tooth loss (trauma vs. caries: OR = 6.51;95% CI:1.45–29.26) and vestibular-lingual position (too vestibular vs. correct: OR = 2.85; 95%CI:1.17–6.93). A sensitivity analysis adding to the final model the presence or not of an implant collar (inferior to 1.8 mm or ≧1.8 mm) did not lead either to a significance of that variable or to the loss of significance of the implant brand. Similarly, no differences were observed when separating in the reference group brand the tissue-level implants from the bone-level ones.

## Discussion

This cross-sectional investigation on a representative sample from patients treated in a university postgraduate clinic has shown that peri-implant diseases are highly prevalent among patients with dental implants. More than 30% of the participants had peri-implantitis, which affected a mean of more than 2 implants in each of those patients. Smoking, moderate/severe periodontitis, having less than 16 remaining teeth, plaque, too vestibular position, implant brand, bridge as restoration type, and trauma as reason of tooth loss were identified as risk indicators of peri-implantitis, while interproximal flossing/brushing, PPIs and anticoagulants as protective ones.

A peculiarity of the data reported in the present study was the use of an intermediate peri-implant health category between peri-implant mucositis and peri-implantitis, denominated pre-periimplantitis. The need to establish this category resulted from the difficulty in identifying initial bone loss in the absence of baseline radiographs. Indeed, it is likely that in such category cases of both peri-implant mucositis and incipient peri-implantitis have been included. This condition was present in 31.7% of the included implants, what highlights the need of having reliable documentation to detect early bone changes and hence, diagnose incipient peri-implantitis, which should be amenable for predictable treatment outcomes. Similarly, if the cases of peri-implant mucositis present bone levels between 1–2 mm, these are probably more amenable for evolving to peri-implantitis and therefore their appropriate management should be a priority.

When comparing the present results with the available literature, these patients demonstrate a slightly higher prevalence and severity of peri-implantitis, counterbalanced by a low prevalence of peri-implant health (Derks & Tomasi, 2015; Derks, Schaller, Hakansson, Wennstrom, Tomasi, & Berglundh, 2016a; Kordbacheh Changi, Finkelstein, & Papapanou, 2019; Rakic et al., 2018; Rodrigo et al., 2018; Romandini et al., 2019; Wada et al., 2019). This finding may be partially related to the characteristics of the present population, which was extrapolated from a university periodontal clinic, resulting in an inferior proportion (27.8%) of participants with no/mild periodontitis when compared to the other studies. Despite this higher prevalence of periodontitis, the present sample was characterized by a scarce control of the disease, being less than 50% the patients with at least one maintenance session/year (Amerio et al., 2020) and 17.9% the plaque-free implants, thus predisposing even more to high peri-implantitis prevalence.

Another aspect that is likely to have influenced the reported prevalence was related to the clinical assessment methods employed. In the present study, BoP was considered positive even in case of one single site out of six presenting with a punctiform drop of blood after deep probing to assess PPD. This strict evaluation method resulted in 91.5% of BoP+ implants, potentially contributing to the high prevalence of peri-implant diseases. We may speculate that, around implants, there are different methods to assess BoP, including profuse vs. punctiform bleeding (Renvert, Persson, Pirih, & Camargo, 2018), or evaluating bleeding after deep probing to assess PPD vs. its evaluation *per sé* by walking marginally with the probe through the peri-implant sulcus. Since these methods have not been standardized among the published studies, this may have contributed to the different reported prevalences in comparison to the present study. The use of a more conservative threshold for peri-implant inflammation (BoP/SoP in at least 4 sites/implant), resulted in 55.2% of the implants presenting with peri-implant health.

Finally, another possible explanation is that the present study included all the implants loaded from at least 1 year which were in the mouth of the selected participants at the time of the examination, resulting in a mean of 4.6 implants/patient. Conversely, most of the studies in the literature only analyzed a subset of implants of the included patients (e.g. the ones placed in the same clinic), leading to a lower number of implants/patient (e.g. 1.7 for Rodrigo, et al., 2018; 2.5 for Kordbacheh Changi, et al., 2019; 3.0 for Wada, et al., 2019; 3.5 for Vignoletti, et al., 2019). This approach prevented in the present study the risk to underestimate the patient-level prevalences of diseases (as, on the contrary, affected implants may be excluded from the analysis in otherwise healthy patients).

The extent of peri-implantitis in patients with > 1 implant was 43.6%. This percentage is similar to the few reports of this epidemiological descriptor which are present in the literature, which ranges from 37.2% of Mir-Mari et al. (Mir-Mari, Mir-Orfila, Figueiredo, Valmaseda-Castellón, & Gay-Escoda, 2012) to 38.2% of Vignoletti et al. (2019), 40.1% of Derks et al. (Derks, Schaller, Hakansson, Wennstrom, Tomasi, & Berglundh, 2016a) and 41.8% of Fransson et al. (Fransson, Wennström, Tomasi, & Berglundh, 2009).

When it comes to risk/protective indicators, the present study was able to identify several factors associated with peri-implantitis. Presence of plaque and the non-use of interproximal hygiene devices were indicators of poor oral hygiene, what has previously reported as a risk indicator with strong evidence for peri-implantitis (Ferreira, Silva, Cortelli, Costa, & Costa, 2006; Schwarz, Derks, Monje, & Wang, 2018). Similarly, the association of the restoration type (bridges vs. single crowns), has also been reported in previous studies (e.g. Rodrigo, et al., 2018), and may be explained by the more difficult access to oral hygiene procedures.

An association with moderate/severe, but not mild, periodontitis was also found, which is in agreement with most of the epidemiological evidence (Derks, Schaller, Hakansson, Wennstrom, Tomasi, & Berglundh, 2016a; Roos-Jansaker, Renvert, Lindahl, & Renvert, 2006; Kordbacheh Changi et al., 2019). The association with the number of remaining teeth could also be interpreted as due to periodontitis, where stage IV cases with extensive tooth loss are characterized by worst peri-implant conditions, and this agrees with reports reporting a different measure of tooth loss (the number of implants) as risk indicator (Derks, et al., 2016a; Vignoletti, et al., 2019).

Regarding the association with smoking, the literature is controversial, with some studies reporting a significant association (Pimentel et al., 2018; Roos-Jansaker et al., 2006), while others not (Dalago, Schuldt Filho, Rodrigues, Renvert, & Bianchini, 2017; Derks, Schaller, Hakansson, Wennstrom, Tomasi, & Berglundh, 2016a). This might be interpreted as a “masking effect” due to periodontitis. However, in the present study, this was not the case and smoking had even a stronger association with peri-implantitis than periodontitis.

Implant brand has been previously reported as a risk indicator for peri-implantitis (Derks, et al., 2016a), which is in agreement with the present findings. While in the study of Derks et al. (2016a) this finding was potentially explainable by the simple presence or not of a collar in the implant design (the reference group was represented by a brand composed at that time only of tissue level implants), in the present study the sensitivity analyses adding to the final model the presence or not of an implant collar, or separating in the reference group the tissue-level implants from the bone-level ones, allowed to discard this hypothesis.

New associations were also found for previously neglected factors. Malposition implants have been considered associated to peri-implant diseases, but previous studies have not systematically studied this factor. In the present study, implants placed too facially accounted for an OR superior than periodontitis, thus supporting this hypothesis. Medications have previously shown to influence implant failures (Chappuis, Avila-Ortiz, Araujo, & Monje, 2018), however there is a paucity of studies analyzing its relation with peri-implant diseases. The use of anticoagulants and of PPIs have resulted as significant protective indicators, what may be explained by their secondary anti-inflammatory effects (Kedika, Souza, & Spechler, 2009; Müller, Chatterjee, Rath, & Geisler, 2015). Finally, trauma as reason for tooth extraction has shown in the present study the highest OR for an association with peri-implantitis. It may be speculated that implants placed after trauma are more frequently placed with type 1 protocols and/or in conjunction with bone augmentation procedures, thus indirectly suggesting a potential role of those factors as risk indicators for peri-implantitis.

Contrarily to other studies, other putative risk indicators were not associated with peri-implantitis in this investigation. Among them, the KTH (Monje & Blasi, 2019; Vignoletti et al., 2019), the retention type (cemented-retained restorations – Kordbacheh Changi et al., 2019; Staubli, Walter, Schmidt, Weiger, & Zitzmann, 2017), the emergence angle and profile (Katafuchi, Weinstein, Leroux, Chen, & Daubert, 2018), the fitness of the prosthesis (Kordbacheh Changi et al., 2019) and a prosthetic design not allowing access to hygiene (Rodrigo, et al., 2018). The final multilevel multivariate model of the present study included plaque and interproximal brushing/flossing around implants, and this may be the reason why those factors were not associated with peri-implantitis, even if KTH and cemented restorations were significantly associated in the univariate analyses. Similarly, clinical signs of occlusal overloading were associated to peri-implantitis in univariate but not in multivariate models, which is supported by previous studies (Schwarz, et al., 2018). Conversely, even if a strong evidence exists for the absence of regular maintenance (Monje et al., 2016; Monje, Wang, & Nart, 2017; Schwarz, Derks, Monje, & Wang, 2018), the present study was not able to highlight this factor as risk indicator for peri-implantitis. This might be due to the self-reported assessment of maintenance frequency, as well as to a higher frequency of maintenance recalls of the most severe periodontitis patients, which are also at increased risk for peri-implantitis.

The present study has reported the prevalence and the risk/protective indicators of peri-implant diseases from a representative sample in a university postgraduate dental clinic, thus minimizing the risk of selection bias. Contrarily to most published data, all implants present in the selected participants (not only the ones placed in the clinic) were analyzed, thus preventing the risk to underestimate the patient-level prevalence of peri-implant diseases. The analysis of many risk/protective indicators of peri-implantitis has allowed to construct a model explaining a great proportion of patient variance. However, this study also has some limitations worth mentioning. Its cross-sectional design doesn’t allow to prove any causality of the identified risk/protective indicators on peri-implantitis. Moreover, as with most of similar studies reported in the literature, the absence of baseline documentation didn’t allow to identify peri-implantitis through direct evidence, what may lead to misclassification bias. Moreover, some of the tested potential risk/protective indicators were not collected with gold standard methods (e.g. self-reported history). Since the sample size calculation was performed based on prevalence data, some of the tested risk/protective indicators may lack the appropriate statistical power to enter in the final model. Another limitation was the impossibility to obtain trusty information regarding previous periimplantitis treatment (Ravidà, Galli, et al., 2020a), loading times and the implant placement surgical protocols employed, which prevented us to include such information in the analyses. Finally, even if representative from a university clinic, the present results suffer from limited generalizability since different prevalence and risk/protective indicators may be found in other populations.

## Conclusions

According to the present study, peri-implant diseases are highly prevalent among patients with dental implants. Several patient-level and implant-level factors were identified as risk and protective indicators of peri-implantitis, which in case of future proof of causality should be included in preventive and therapeutic strategies. Consequently, randomized clinical trials or, when not possible for ethical reasons, prospective cohort studies are needed to demonstrate true causality of the identified risk/protective indicators and to study the preventive efficacy of their modification.

## Data Availability

Upon request to the corresponding author, the data could be shared

## Acknowledgments

The authors wish to kindly thank Mariano del Canto, Laura Sobrino and Javier Calatrava for their kind support during part of the data collection process.

The authors declare no conflicts of interest related to this study. This study was partially funded by the Osteology Foundation through a Young Research Grant to Dr. Mario Romandini (project n. 15–251).

## Author contributions

M.R. conceived the ideas; C.L., I.P., A.A., M.C.S. and M.R. collected the data; M.R., M.C.S. and M.S. analysed the data; and M.R. and M.S. led the writing.

## Supporting Information

Additional Supporting Information may be found in the online version of this article:

**Table S1.** Distribution of the putative patient-level risk/protective indicators in the study population, overall and according to peri-implantitis status.

**Table S2.** Distribution of the putative implant-level risk/protective indicators in the study implants, overall and according to peri-implantitis status.

**Table S3.** Risk/protective indicators associated in univariate analyses with peri-implantitis: patient-level variables.

**Table S4.** Risk/protective indicators associated in univariate analyses with peri-implantitis: implant-level variables.

## Notes

### Competing Interest Statement

The authors have declared no competing interest.

### Author Declarations

The research protocol was ethically approved by the CEIC Hospital Clinico San Carlos, Madrid, Spain (19/182-E).

